# Cause-specific mortality across ICD10 chapters in steatotic liver disease stratified by subtypes and fibrosis status

**DOI:** 10.64898/2025.12.23.25342903

**Authors:** Qi Feng, Pinelopi Manousou, Chioma N Izzi-Engbeaya, Mark Woodward

## Abstract

**Background and aims:** To examine the associations between Steatotic liver disease (SLD), its subtypes (MASLD, MetALD, ALD), fibrosis stages and mortality from a broad spectrum of diseases.

**Methods:** We analysed 486156 UK Biobank participants. We summarised the causes of death during follow-up, by SLD status and subtypes. Multivariable Cox models estimated associations between SLD, SLD subtypes, FIB4 score, and cause-specific mortality outcomes across ICD10 disease categories.

**Results:** Among 178336 participants with SLD, 20766 died over a median follow up of 13.8 years. The leading causes of deaths were neoplasm (46.4%), circulatory disease (24.2%), respiratory disease (7.0%), and digestive disease (4.8%). Compared to participants without SLD (n=307820), those with SLD had significantly higher mortality from neoplasm (HR 1.24 (1.20, 1.28)), circulatory (1.57 (1.50, 1.64)), and digestive disease (1.85 (1.67, 2.05)), as well as from metabolic, and genitourinary diseases, across all FIB4 score levels. Elevated risks were also observed for mortality from diabetes (3.40 (2.59, 4.46))), Covid-19 (1.97 (1.76, 2.21)), myocardial infarction (1.69 (1.53, 1.86)), stroke (1.24 (1.04, 1.48)), and several cancers, such as breast (1.50 (1.35, 1.68)), colorectal (1.22 (1.11, 1.34)), and prostate cancer (1.22 (1.09, 1.37)). SLD subtypes exhibited distinct patterns of cause-specific mortality.

**Conclusion:** SLD is associated with increased mortality across a wide range of disease categories, with extrahepatic cancers and circulatory diseases being the primary contributors. These findings underscore the importance of multidisciplinary management strategies targeting liver fibrosis, cardiometabolic burden, and cancer prevention in individuals with SLD.

**Graphic abstract:** 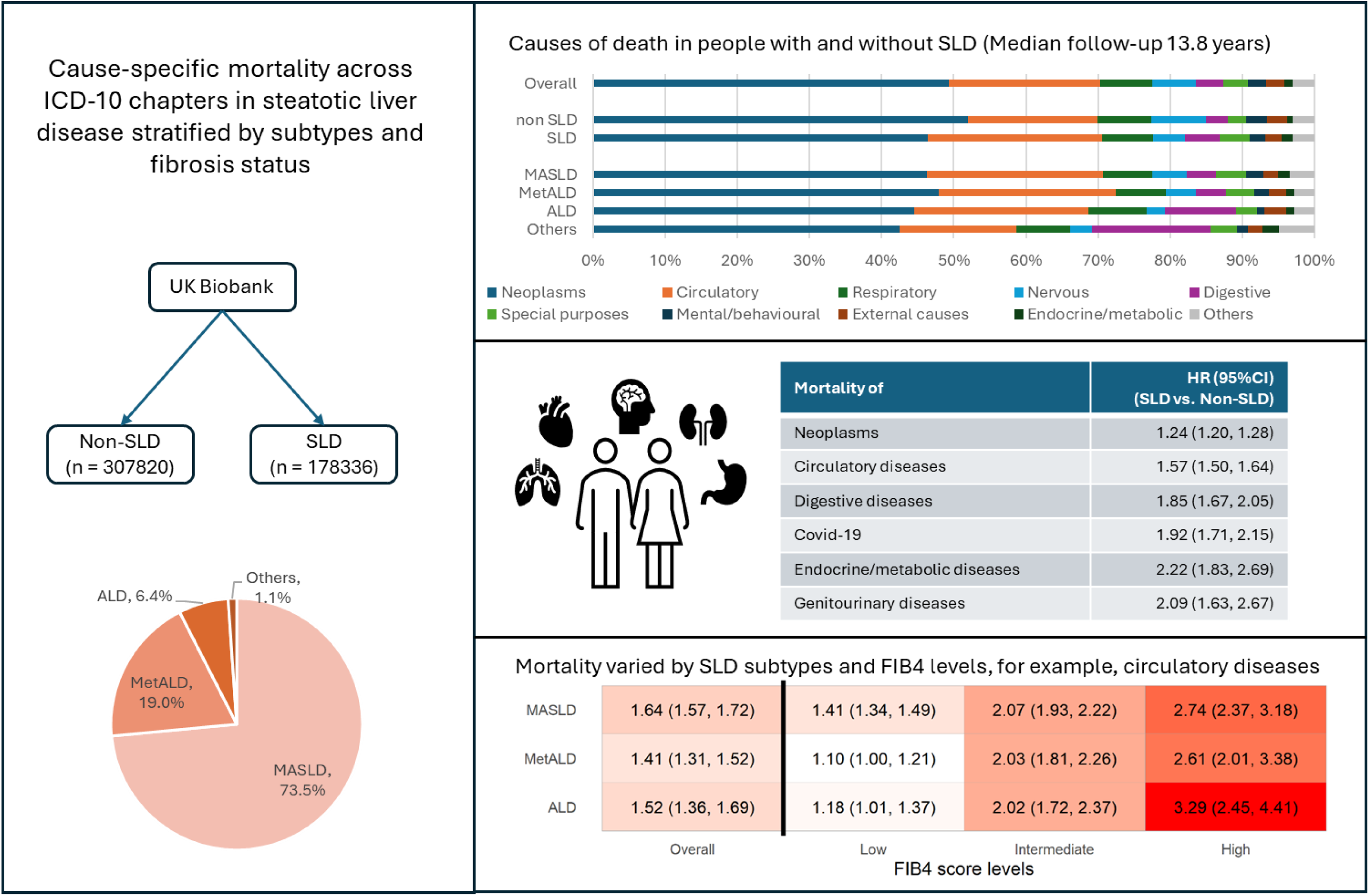

## 1. Introduction

Steatotic liver disease (SLD) is the most common chronic liver condition, affecting one in three adults globally ^1^. It encompasses three subtypes: metabolic dysfunction associated steatotic liver disease (MASLD), alcohol related liver disease (ALD), and metabolic dysfunction and alcohol-related steatotic liver disease (MetALD). MASLD is linked to increased risk of various extrahepatic conditions, including cardiovascular disease (CVD), and extrahepatic cancers ^2,3^.

Despite increasing recognition of its systemic implications, evidence on impacts of SLD on long-term mortality remains inconsistent. Most prior studies concentrated on a limited set of outcomes (e.g., all-cause mortality, liver related mortality, CVD and cancer mortality), with mixed results. A systematic review reported that MASLD increased mortality overall and from liver-related diseases, but not from CVD or cancers ^4^. Another meta-analysis reported null associations with all-cause mortality and liver related mortality ^5^. These discrepancies may be due to variation in study populations, and disease severity.

Several important gaps remain. First, most existing research has focused narrowly on a few causes of death, leaving many other potential causes underexplored. Second, comparative evidence on mortality risk across the three SLD subtypes is sparse, limiting our understanding of subtype-specific prognosis. To address these limitations, this study aimed to provide a comprehensive assessment of mortality patterns in individuals with SLD using a large UK database. We examined all major causes of death across the ICD10 categories, and assessed their associations with SLD, subtypes, and fibrosis.

## 2. Methods

### 2.1 Data and participants

This study utilised the UK Biobank data, a prospective cohort comprising 500000 UK residents aged 40 - 70 years, recruited 2006-2010. Baseline assessment included socioeconomic status, lifestyle, environmental exposure, medical history, and physical measures. Participants were followed up via linkage to national death registries, and hospital records.

SLD was defined as fatty liver index (FLI) ≥ 60. FLI is an indicator for liver steatosis, ^6^. The cutoff value of 60 to define liver steatosis has been used extensively and validated ^7^. In a sensitivity analysis, we replaced FLI with hepatic steatosis index (HSI), another measure for liver steatosis ^8^. SLD subtypes (MASLD, MetALD, and ALD) were defined based on alcohol consumption and presence of cardiometabolic risk factors (CMRFs), using the definition recommended by the latest SLD guideline ^9^. The daily alcohol consumption cutoff values for MASLD, MetALD and ALD were < 20/30 (females/males), 20-50/30-60, and > 50/60 g/day, respectively. We excluded women who were pregnant at baseline, and people who had missing data for calculating FLI.

Participants with presence of liver steatosis were classified as having MASLD, MetALD or ALD, and as Others if they had viral hepatitis, hemochromatosis, Wilson’s disease, biliary cirrhosis, autoimmune hepatitis, primary sclerosing cholangitis, drug-induced liver injuries or Budd-Chiari syndrome. These chronic liver diseases were ascertained via hospital records; the code lists for these conditions are presented in supplementary methods.

Participants with SLD were further categorised into three groups based on FIB4 score, a non-invasive marker for fibrosis status. We used the cutoff values 1.30 and 2.67 to categorise low, intermediate and high levels of FIB-4 score for people < 65 years, and 2.00 and 2.67 for people ≥ 65 years ^10^.

### 2.2 Covariates

Ethnicity was classified into White, and mixed/others. The Townsend Deprivation Index was used to assess socioeconomic status. Lifestyle factors included smoking status (current, previous and never smoker), alcohol intake, and physical activity level. Alcohol intake was assessed via self-reported consumptions of various alcoholic drinks summed up to derive the average alcohol consumption (g/day). Physical activity level was categorized into low, moderate and high levels, based on the frequency, duration and intensity of their physical activities. Systolic and diastolic blood pressure (BP) were measured twice and the averages were used in analyses. Blood biochemistry markers were measured at the central laboratory, including triglycerides (TG), high density lipoprotein (HDL), and glycated haemoglobin (HbA1c). Aligning with the SLD guideline, we considered five CMRFs: Overweight/obesity was defined as BMI ≥25 kg/m² or waist circumference >94 cm (male) or >90 cm (female). Prediabetes/diabetes was defined as HbA1c ≥48mmol/mol or diagnosis of type 2 diabetes or on treatment for type 2 diabetes. Hypertension was defined as systolic BP ≥ 130 or diastolic BP ≥ 85 mmHg or on antihypertensive drug treatment or diagnosis of hypertension. High TG was defined as plasma TG ≥1.70 mmol/L or on lipid lowering treatment. Low HDL was defined as HDL-cholesterol ≤1.0 mmol/L (male) or ≤1.3 mmol/L (female) or on lipid lowering treatment^9^.

### 2.3 Outcomes

We examined causes of death by ICD10 disease category, including infectious/parasitic (ICD10 codes: A00-B99), neoplasms (C00-D48), haematological (D50-D89), endocrine/ metabolic (E00-E99), mental/behavioral (F00-F99), nervous system (G00-G99), eye/adnexa (H00-H59), ear/mastoid (H60-H69), circulatory (I00-I99), respiratory (J00-J99), digestive (K00-K99), skin/subcutaneous (L00-L99), musculoskeletal (M00-M99), genitourinary (N00-N99), pregnancy/childbirth (O00-O99), congenital (Q00-Q99), symptoms/signs (R00-R99), injury/poisoning (S00-T98), external causes (V01-Y98), special purpose codes (U00-U99) and health services (Z00-Z99). We additionally evaluated the associations with common disease-specific causes in this cohort. Participants were censored at the date of death or the last date of follow-up (30 November 2022), whichever occurred first.

### 2.4 Statistical analysis

We summarised the number of deaths from each ICD10 disease category, and identified the top 10 most common ICD10 codes in each category. Cox models were used to assess the association between SLD, subtypes, FIB4 score and mortality, expressed as hazard ratio (HR) and 95% confidence interval (CI), using individuals without SLD as reference. Models were stratified by region and age group (< vs. ≥ 65 years old), and adjusted for sex, ethnicity, education, Townsend Deprivation Index (in fifths), physical activity, smoking, and daily alcohol consumption. Cox models were not fitted for outcomes with < 50 deaths. The proportional hazard assumption was examined by scaled Schoenfeld residuals, and no evidence was observed for violation.

We conducted several sensitivity analyses. First, we additionally adjusted for the CMRFs to examine whether the associations were independent of these risk factors. Second, we excluded the first two years of follow-up to allow for reverse causation bias. Third, alternative to FLI, we used HSI as a surrogate for liver steatosis, with HSI > 36 indicating presence of liver steatosis. Although a subset of UK Biobank participants underwent liver MRI scans, which was a more accurate measure for liver fat, we could not conduct a similar sensitivity analysis, because FIB4 data were not available at the time of the MRI scans. Fourth, we fitted Fine-Gray model to examine the associations, to account for competing risks to cause-specific death. We also conducted sex-specific analyses.

## 3. Results

Among 486156 eligible participants, 178836 were identified as having SLD (age 57.3 years, 36.3% females). Among people with SLD, 73.5% had MASLD, 19.0% MetALD, and 6.4% ALD (figure 1). Individuals with SLD were more likely to be older, male, more socioeconomically deprived, less educated, less physically active, and to have higher alcohol consumption, BMI, waist circumference, BP, TG, HbA1c, and to have lower HDL, compared to those without SLD. (table 1; supplementary Table 1)

**Figure 1:**
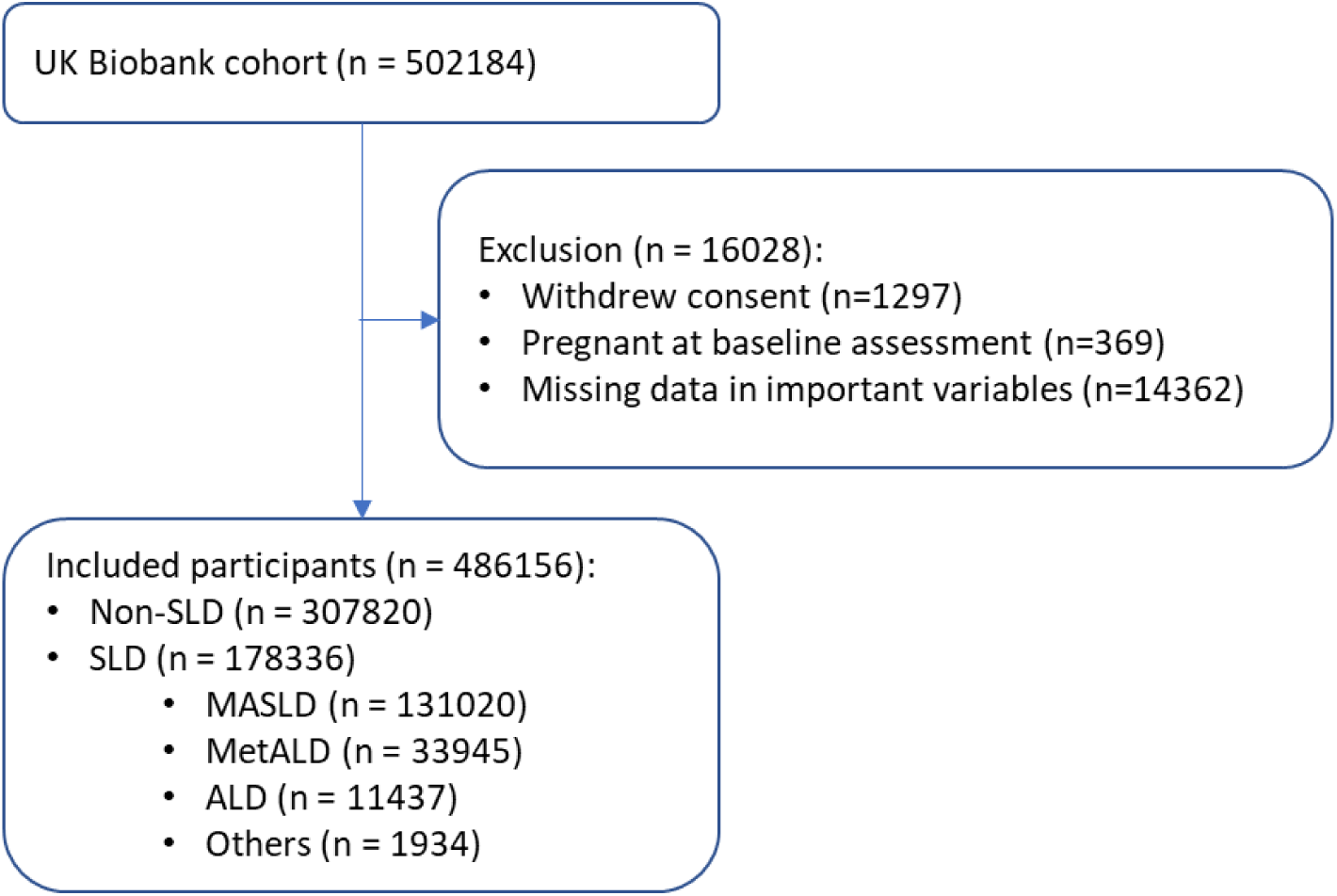
flowchart of participant selection. SLD: steatotic liver disease. MASLD: metabolic dysfunction associated steatotic liver disease. MetALD: metabolic dysfunction and alcohol related liver disease. ALD: alcohol related liver disease.

**Table 1:** Baseline characteristics of people with and without SLD in the UK Biobank.

|  | Non-SLD | SLD |  |  |  | Overall |
| --- | --- | --- | --- | --- | --- | --- |
|  |  | All SLD | MASLD | MetALD | ALD |  |
|  | n = 307820 | n = 178336 | n = 131020 | n = 33945 | n = 11437 | n = 486156 |
| Sex, male | 108955 (35.4%) | 113587 (63.7%) | 76275 (58.2%) | 25701 (75.7%) | 10312 (90.2%) | 222542 (45.8%) |
| Age, years | 56.1 (8.2) | 57.3 (7.8) | 57.4 (7.9) | 57.0 (7.7) | 56.3 (7.7) | 56.6 (8.1) |
| Townsend deprivation index |  |  |  |  |  |  |
| 1st fifth (least deprived) | 65121 (21.2%) | 32462 (18.2%) | 23791 (18.2%) | 6619 (19.5%) | 1799 (15.7%) | 97583 (20.1%) |
| 5th fifth (most deprived) | 56140 (18.2%) | 40870 (22.9%) | 30641 (23.4%) | 6584 (19.4%) | 2950 (25.8%) | 97010 (20.0%) |
| Education, higher education | 109872 (35.7%) | 47346 (26.5%) | 34208 (26.1%) | 9794 (28.9%) | 2811 (24.6%) | 157218 (32.3%) |
| Ethnicity, White | 290659 (94.4%) | 168183 (94.3%) | 121919 (93.1%) | 33225 (97.9%) | 11215 (98.1%) | 458842 (94.4%) |
| Smoking, never | 179488 (58.3%) | 85523 (48%) | 69146 (52.8%) | 12397 (36.5%) | 3232 (28.3%) | 265011 (54.5%) |
| Alcohol drinking, g/d* | 9.7 (0.7, 20.0) | 11.9 (0.0, 28.6) | 5.7 (0.0, 14.8) | 37.9 (32.2, 45.9) | 75.4 (65.8, 91.6) | 10.2 (0.4, 22.4) |
| Physical activity, high | 106203 (34.5%) | 47835 (26.8%) | 33406 (25.5%) | 10236 (30.2%) | 3703 (32.4%) | 154038 (31.7%) |
| Hypertension | 188397 (61.2%) | 145417 (81.5%) | 105177 (80.3%) | 28663 (84.4%) | 10115 (88.4%) | 333814 (68.7%) |
| Overweight/obesity | 172114 (55.9%) | 175794 (98.6%) | 129553 (98.9%) | 33341 (98.2%) | 11058 (96.7%) | 347908 (71.6%) |
| Prediabetes/diabetes | 38433 (12.5%) | 51762 (29.0%) | 40909 (31.2%) | 7641 (22.5%) | 2595 (22.7%) | 90195 (18.6%) |
| High triglycerides | 77920 (25.3%) | 128292 (71.9%) | 95116 (72.6%) | 23904 (70.4%) | 8057 (70.4%) | 206212 (42.4%) |
| Low HDL-cholesterol | 92556 (30.1%) | 73177 (41.0%) | 61157 (46.7%) | 9109 (26.8%) | 2117 (18.5%) | 165733 (34.1%) |
| FIB4 score level |  |  |  |  |  |  |
| Low | NA | 119362 (66.9%) | 90035 (68.7%) | 21825 (64.3%) | 6425 (56.2%) | NA |
| Intermediate | NA | 49888 (28.0%) | 34669 (26.5%) | 10465 (30.8%) | 4162 (36.4%) | NA |
| High | NA | 4461 (2.5%) | 2766 (2.1%) | 861 (2.5%) | 610 (5.3%) | NA |
| Unknown | NA | 4625 (2.6%) | 3550 (2.7%) | 794 (2.3%) | 240 (2.1%) | NA |

SLD: steatotic liver disease. MASLD: metabolic dysfunction associated steatotic liver disease. MetALD: metabolic dysfunction and alcohol related liver disease. ALD: alcohol related liver disease. HDL: high density lipoprotein. The cutoff values for low, intermediate and high FIB4 scores were 1.30 and 2.67 for people < 65 years old, and 2.00 and 2.67 for people ≥ 65 years old. \*: showing median(interquartile interval)

FIB4 scores were not available for 4625 (2.6%) participants with SLD. Among those with available FIB4 scores, 68.7% had low FIB4 scores, while 28.7% and 2.6% had intermediate and high scores, respectively.

### 3.1 Causes of death

During a median follow-up of 13.8 years, 42520 deaths were recorded: 20766 (48.8%) in participants with SLD. Overall, six ICD10 disease categories accounted for 90% of all deaths: neoplasms (49.3%), circulatory (21.0%), respiratory (7.2%), nervous (6.0%), digestive (3.9%) diseases and special purpose codes (3.3%). Compared to individuals without SLD, those with SLD had a higher proportion of deaths from circulatory (24.2% vs. 17.9%), special purpose codes (4.1% vs. 2.5%), endocrine/metabolic (1.5% vs. 0.9%) and digestive (4.8% vs. 3.1%) diseases. (figure 2, supplementary table 2)

**Figure 2:**
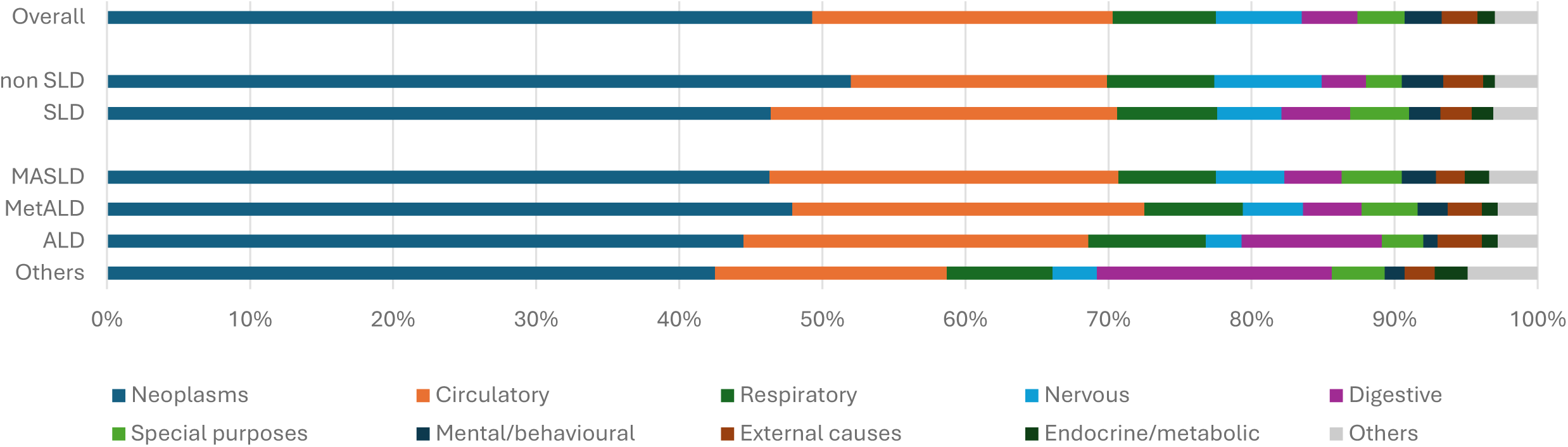
Causes of death in people with and without SLD. The numbers show the percentage. The cutoff values for low, intermediate and high FIB4 scores were 1.30 and 2.67 for people < 65 years old, and 2.00 and 2.67 for people >= 65 years old. SLD: steatotic liver disease. MASLD: metabolic dysfunction associated steatotic liver disease. MetALD: metabolic dysfunction and alcohol related liver disease. ALD: alcohol related liver disease. The ICD10 chapters were defined using the following ICD10 codes: neoplasms (C00-D48), circulatory (I00-I99), respiratory (J00-J99), nervous system (G00-G99), digestive (K00-K99), special purpose codes (U00-U99) mental/behavioral (F00-F99), external causes (V01-Y98), endocrine/ metabolic (E00-E99).

Among neoplasms-related deaths, the most common cancers were lung (C34, 16.9%), pancreatic (C25, 7.5%), breast (C50, 7.3%), prostate (C61, 6.0%) and colon (C18, 5.3%) (supplementary table 3). Liver cancer (C22) accounted for 3.4% of neoplasm-related death (2.6% and 4.1% in people without and with SLD, respectively). Leading causes of circulatory mortality were ischemic heart disease (I25, 30.3%), myocardial infarction (I21, 20.4%), and stroke (I64, 6.4%). For respiratory disease, the top contributor was COPD (J44, 37.2%). Alzheimer’s disease (G30, 31.6%) and Parkinson’s disease (G20, 21.6%) were the top causes of nervous system deaths. Nearly all deaths in the special purpose codes chapter were attributed to Covid-19 (U07, 97.3%). In digestive disease category, ALD (K70, 20.1%), vascular disorders of intestine (K55, 11.2%) and hepatic fibrosis/cirrhosis (K74, 9.1%) were the most common; overall 66.6% were liver-related (K70-K77). Dementia (F03, 66.0%; F01, 28.7%) and diabetes (E11, 30.6%; E14, 23.5%) were the most common mental/behavioral and metabolic causes, respectively. Chronic kidney disease (N18) contributed 19.4% of the genitourinary deaths.

### 3.2 Associations with mortality outcomes

Overall mortality was higher in people with SLD compared to those without, with an absolute mortality rate difference of 3.53/1000 person-years (8.78 vs. 5.25 /1000 person-years), corresponding to a 28% higher mortality (HR (95%CI): 1.28 (1.26, 1.31)). Mortality rates increased progressively with higher FIB4 score: compared to those without SLD, participants with SLD with low, intermediate and high FIB4 scores also had higher mortality by 11% (1.11 (1.09, 1.14)), 63% (1.63 (1.58, 1.68)), and 162% (2.62 (2.48, 2.77)), respectively.

By ICD10 disease categories, SLD was significantly associated with mortality from neoplasms (SLD vs non-SLD: 1.24 (1.20, 1.28)), circulatory diseases (1.57 (1.50, 1.64)), digestive diseases (1.85 (1.67, 2.05)), special purpose codes (1.92 (1.71, 2.15)), endocrine/metabolic disease (2.22 (1.83, 2.69)), genitourinary disease (2.09 (1.63, 2.67)) and skin/subcutaneous diseases (3.11 (1.83, 5.19)) (table 2). Furthermore, mortality from respiratory diseases for intermediate and high FIB4 scores vs. non-SLD showed HRs of 1.27 (1.14, 1.43) and 1.61 (1.28, 2.03), respectively. We found no evidence for increased mortality from infectious/parasitic, mental/behavioural, musculoskeletal, nervous system diseases or external causes.

**Table 2:** Adjusted associations between SLD, FIB4 scores and cause-specific mortality.

|  |  | SLD |  |  |  |  |
| --- | --- | --- | --- | --- | --- | --- |
|  | Non-SLD<br>n = 307820 | All SLD<br>n = 178336 | Low FIB4<br>n = 119362 | Intermediate FIB4<br>n = 49888 | High FIB4<br>n = 4461 | P for trend |
| **Neoplasms (C00-D48)** |  |  |  |  |  |  |
| Death, n | 11239 | 9564 | 5981 | 2819 | 512 |  |
| Mortality rate, /1000py | 2.71 | 4.05 | 3.75 | 4.3 | 9.6 |  |
| HR (95%CI) | Reference | 1.24 (1.20, 1.28) | 1.11 (1.08, 1.15) | 1.50 (1.44, 1.57) | 2.22 (2.02, 2.42) | < 0.01 |
| **Circulatory (I00-I99)** |  |  |  |  |  |  |
| Death, n | 3860 | 4997 | 2977 | 1575 | 306 |  |
| Mortality rate, /1000py | 0.93 | 2.11 | 1.87 | 2.4 | 5.74 |  |
| HR (95%CI) | Reference | 1.57 (1.50, 1.64) | 1.34 (1.28, 1.41) | 2.08 (1.96, 2.22) | 2.88 (2.56, 3.25) | < 0.01 |
| **Digestive (K00-K99)** |  |  |  |  |  |  |
| Death, n | 676 | 988 | 417 | 325 | 217 |  |
| Mortality rate, /1000py | 0.16 | 0.42 | 0.26 | 0.5 | 4.07 |  |
| HR (95%CI) | Reference | 1.85 (1.67, 2.05) | 1.15 (1.01, 1.30) | 2.55 (2.21, 2.93) | 13.97 (11.87, 16.44) | < 0.01 |
| **Special purpose codes (U00-U99)** |  |  |  |  |  |  |
| Death, n | 549 | 837 | 509 | 269 | 38 |  |
| Mortality rate, /1000py | 0.13 | 0.35 | 0.32 | 0.41 | 0.71 |  |
| HR (95%CI) | Reference | 1.92 (1.71, 2.15) | 1.66 (1.46, 1.88) | 2.65 (2.27, 3.09) | 2.59 (1.86, 3.62) | < 0.01 |
| **Endocrine/metabolic (E00-E99)** |  |  |  |  |  |  |
| Death, n | 179 | 315 | 180 | 99 | 20 |  |
| Mortality rate, /1000py | 0.04 | 0.13 | 0.11 | 0.15 | 0.37 |  |
| HR (95%CI) | Reference | 2.22 (1.83, 2.69) | 1.82 (1.47, 2.26) | 2.92 (2.26, 3.79) | 4.91 (3.06, 7.89) | < 0.01 |
| **Genitourinary (N00-N99)** |  |  |  |  |  |  |
| Death, n | 111 | 188 | 103 | 54 | 21 |  |
| Mortality rate, /1000py | 0.03 | 0.08 | 0.06 | 0.08 | 0.39 |  |
| HR (95%CI) | Reference | 2.09 (1.63, 2.67) | 1.61 (1.22, 2.12) | 2.78 (1.97, 3.92) | 7.30 (4.50, 11.84) | < 0.01 |
| **Skin/subcutaneous (L00-L99)** |  |  |  |  |  |  |
| Death, n | 22 | 48 | 23 | 17 | 7 |  |
| Mortality rate, /1000py | 0.01 | 0.02 | 0.01 | 0.03 | 0.13 |  |
| HR (95%CI) | Reference | 3.11 (1.83, 5.29) | 2.05 (1.12, 3.76) | 6.07 (3.09, 11.93) | 15.75 (6.43, 38.60) | < 0.01 |
| **Respiratory (J00-J99)** |  |  |  |  |  |  |
| Death, n | 1611 | 1438 | 908 | 406 | 79 |  |
| Mortality rate, /1000py | 0.39 | 0.61 | 0.57 | 0.62 | 1.48 |  |
| HR (95%CI) | Reference | 1.01 (0.94, 1.09) | 0.89 (0.82, 0.97) | 1.27 (1.14, 1.43) | 1.61 (1.28, 2.03) | < 0.01 |

HR (95%CI): hazard ratio (95% confidence interval). py: person-years. SLD: steatotic liver disease. Model was stratified by region and age group (<65 vs. ≥65) and adjusted for sex, ethnicity, education, Townsend Deprivation Index, physical activity level, smoking, and drinking. The cutoff values for low, intermediate and high FIB4 scores were 1.30 and 2.67 for people < 65 years old, and 2.00 and 2.67 for people ≥ 65 years old.

### 3.3 Associations by SLD subtypes

All three SLD subtypes were consistently associated with mortality from neoplasms, circulatory, digestive diseases and special purpose codes, although effect sizes varied. (figure 3)

**Figure 3:**
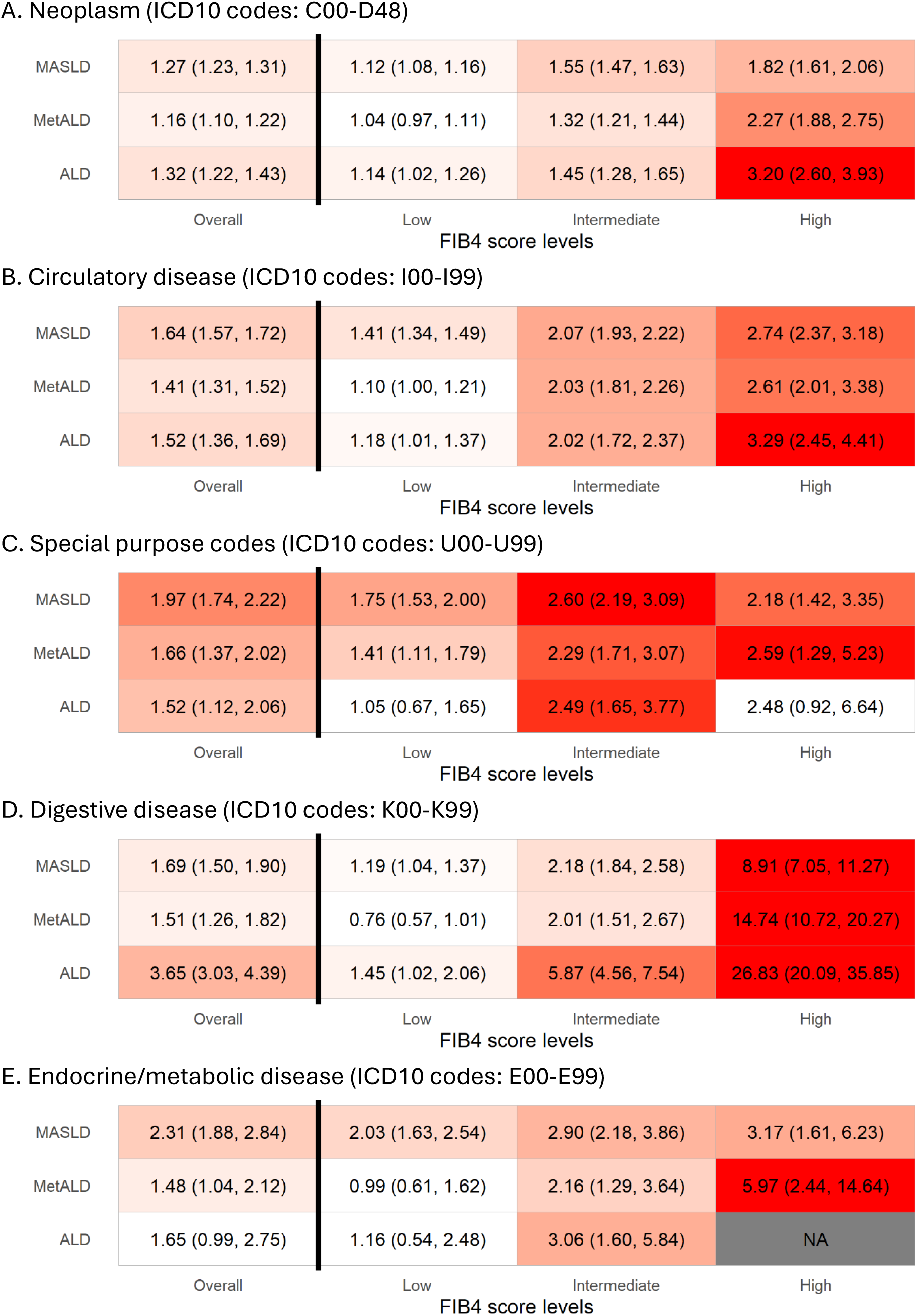
Associations between SLD subtypes, FIB4 score levels and mortality of neoplasm, circulatory disease, Covid-19, digestive diseases and endocrine diseases. The figure shows hazard ratio (95% confidence interval). Model was stratified by region and age group (<65 vs. >=65) and adjusted for sex, ethnicity, education, Townsend Deprivation Index, physical activity level, smoking, and drinking. MASLD: metabolic-dysfunction associated steatotic liver disease. MetALD: metabolic and alcohol associated steatotic liver disease. ALD: alcohol related liver disease

MASLD was associated with the highest HR for endocrine/metabolic disease mortality (2.31 (1.88, 2.84)), higher than MetALD (1.48 (1.04, 2.12)) and ALD (1.65 (0.99, 2.75)). MASLD also showed highest HR with circulatory disease mortality (1.64 (1.57, 1.72)), higher than MetALD (1.41 (1.31, 1.52)) and ALD (1.52 (1.36, 1.69)). It also showed the highest risk for Covid-19-dominated special purpose related mortality (1.97 (1.74, 2.22)).

ALD had the strongest association with digestive disease mortality (3.65 (3.03, 4.39)), higher than MASLD (1.69 (1.50, 1.90)) and MetALD (1.51 (1.26, 1.82)). ALD also showed a similarly elevated cancer mortality risk to MASLD (1.32 (1.22, 1.43) vs. 1.27 (1.23, 1.31)), higher than MetALD (1.16 (1.10, 1.22)).

MetALD was associated with increased mortality across these categories, but typically with lower HR than MASLD or ALD, suggesting an intermediate risk profile.

FIB4 score levels showed a consistent dose-response relationship with each cause-specific mortality. Genitourinary disease mortality was associated with MASLD (2.26 (1.74, 2.93)), but not with MetALD (1.30 (0.81, 2.09)) or ALD (1.50 (0.77, 2.92)) (supplementary figure 1).

### 3.4 Sensitivity analyses

Sensitivity analyses adjusting for CMRFs and excluding the first two years of follow-up showed similar results to the primary analysis. Sensitivity analysis replacing FLI with HSI for liver steatosis also yielded consistent findings. Among all participants with available data on FLI and HSI, the FLI definition identified 178336 participants with SLD, while the HSI definition identified 211097 SLD cases, including 163467 (77.4%) MASLD, 35850 MetALD (17.0%) and 9359 (4.4%) SLD. The findings closely mirror the primary results using FLI, reinforcing the robustness of our conclusion across different case definitions. For example, when examining the association between SLD subtypes and mortality from neoplasm, the HR estimates were 1.17 (1.14, 1.21), 1.09 (1.03, 1.15), and 1.19 (1.09, 1.30) for MASLD, MetALD and ALD, respectively; for circulatory disease mortality, the HR estimates were 1.58 (1.51, 1.65), 1.33 (1.23, 1.44) and 1.42 (1.25, 1.60). (supplementary table 4).

To account for competing risk, we fitted Fine-Gray models to assess the associations between SLD subtypes and cause-specific mortality, showing overall consistent results with the primary analysis. For mortality from neoplasm, the HR estimates were 1.26 (1.22, 1.30), 1.16 (1.10, 1.22) and 1.30 (1.21, 1.39) for MASLD, MetALD and ALD, respectively. For mortality from circulatory diseases, the according HR estimates were 1.64 (1.57, 1.72), 1.41 (1.31, 1.51) and 1.48 (1.33, 1.66) (Supplementary table 5). For other outcomes with smaller number of events, the direct of the associations remained although the 95%CI turned out to be wide, which is a common problem in Fine-Gray model.

Positive associations between SLD subtypes and mortality from neoplasms, endocrine/metabolic, circulatory, digestive and respiratory diseases were observed in both females and males (Supplementary table 6). However, the associations for MASLD were generally stronger in females than in males, particularly for mortality from neoplasm, endocrine/metabolic, circulatory, digestive, respiratory diseases and Covid-19.

### 3.5 Associations with mortality from specific diseases

We observed positive associations of SLD with mortality from female breast cancer (C50, HR 1.50 (1.35, 1.68)), male prostate cancer (C61, 1.22 (1.09, 1.37)), colorectal cancer (C18-C21, 1.22 (1.11, 1.34)), pancreatic cancer (C25, 1.33 (1.20, 1.48)), ischemic heart disease (I25, 1.73 (1.59, 1.87)), myocardial infarction (I21, 1.69 (1.53, 1.86)), stroke (I64, 1.24 (1.04, 1.48)), diabetes (E10-E14, 3.40 (2.59, 4.46)), and Covid-19 (U07, 1.97 (1.76, 2.21)). There was a marginal association between MASLD and lung cancer mortality (1.09 (1.01, 1.17)). We observed negative associations with mortality of COPD, Alzheimer’s disease, and Parkinson’s disease. (table 3)

**Table 3:**
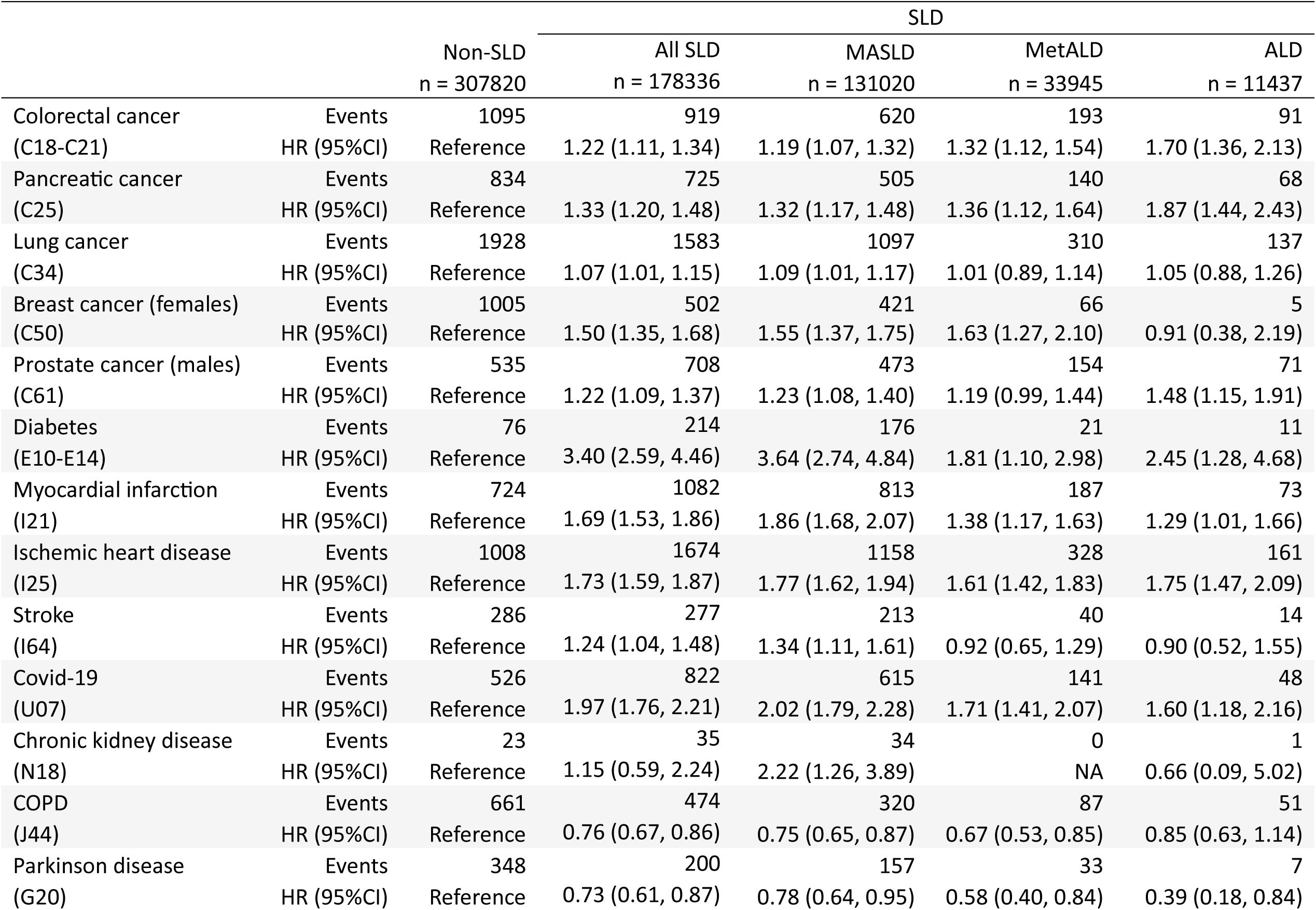
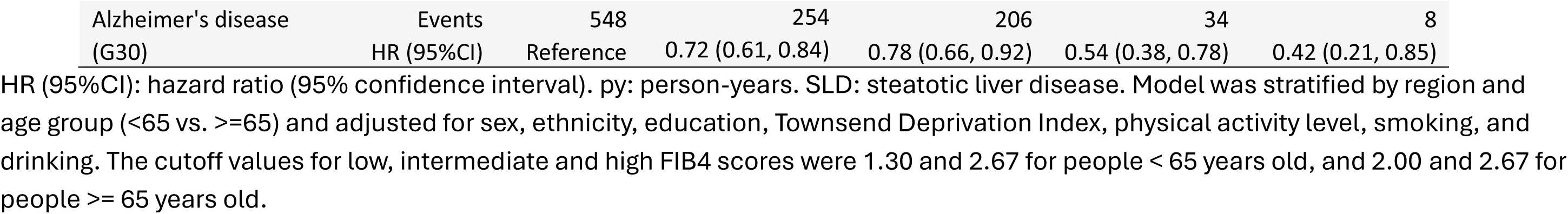
Adjusted associations between SLD, SLD subtypes and selected disease-specific mortality.

## 4. Discussion

In this UK-based cohort, neoplasm and circulatory diseases were the predominant causes of death, jointly representing over 70% of all deaths, whereas deaths from digestive disease accounted for a small proportion (4.8%), and 66.6% of these deaths were due to liver diseases. SLD were associated with elevated mortality of neoplasm, circulatory disease, digestive disease, endocrine/metabolic diseases, Covid-19, and genitourinary disease.

Although neoplasm and circulatory disease were the leading causes of death across all SLD subtypes, consistent with previous findings ^11,12^, SLD subtypes demonstrated distinct cause-specific mortality patterns. Particularly, individuals with ALD showed a substantially higher proportion of deaths from digestive diseases than MASLD and MetALD. Among all digestive disease related death, liver-related conditions, including ALD itself and fibrosis/cirrhosis, accounted for the majority across all subtypes: 58.2%, 86.1% and 79.1% for MASLD, MetALD and ALD, respectively, corresponding to the overall liver related mortality of 2.3%, 3.5% and 7.8%. These findings highlighted the disproportionate burden of liver-related mortality in individuals with ALD.

SLD showed elevated mortality from several major disease categories, including neoplasm, circulatory disease, digestive disease, and endocrine/metabolic disease. These findings are consistent with previous evidence. For example, Simon *et al.* found MASLD was associated with higher mortality from neoplasm, circulatory disease, liver cirrhosis, and hepatocellular carcinoma ^13^, echoing the findings of a Korean cohort study^14^. We found that ALD had strongest associations with digestive-related mortality (mainly liver-related deaths), consistent with a Swedish study where ALD showed the strongest association with liver decompensation ^15^. This may be partly explained by the higher prevalence of advanced fibrosis among people with ALD ^16^. In contrast, MASLD exhibited stronger associations with mortality from circulatory and endocrine/metabolic diseases, reflecting the higher burden of cardiometabolic burdens in MASLD ^17^.

Beyond these commonly investigated outcomes, we also found evidence linking SLD and mortality from skin/subcutaneous, genitourinary and respiratory diseases. Due to the low number of events skin/subcutaneous disease related deaths, we were unable to assess associations with specific ICD10 codes in this category. In the genitourinary category, MASLD showed strong associations with chronic kidney disease, the predominant diagnosis in this category, in line with previous studies ^18^. For respiratory disease mortality overall, only intermediate and high FIB4 scores were linked to higher risk, similar to previous findings ^19^. However, MASLD and MetALD both were associated with lower mortality of COPD, the most frequent respiratory cause of death. Although MASLD has been reported as an independent risk factor for COPD, particularly among men in a Japanese cohort ^20^, its association with COPD-specific mortality has not been well established, and future investigations are warranted.

We also found that SLD was associated with Covid-19 mortality, and MASLD showed the strongest associations, suggesting that the cardiometabolic burdens may exacerbate outcomes in covid-19 infection. Several studies ^21,22^ similarly reported slightly higher Covid-19 mortality in individuals with NAFLD than people without NAFLD, but the difference was not significant, likely due to small samples. A previous review of 5198 individuals found that NAFLD increased the risk of severe Covid-19 by 67%, but did not significantly affect Covid-19 mortality^23^. Our study provided evidence for such an association with a much larger sample size. Hoffmann et al. supported a bidirectional positive association between Covid-19 infection and NAFLD, with Covid-19 aggravating NAFLD, and NAFLD increasing the risk of severe Covid-19, which is attributed to higher infection susceptibility, impaired immune response, increased risk of coagulation and presence of comorbidities ^24^.

Our findings were generally consistent with a Swedish nationwide cohort ^19^, which examined cause-specific mortality in 13000 individuals with clinical diagnosed MASLD. That study reported increased mortality from cancers, circulatory, digestive, respiratory, mental health and endocrine disorders. However, several important differences distinguish our work. First, the Swedish cohort identified MASLD cases via ICD10 codes, which preferentially identified severe cases. Second, their adjustment for confounding was limited to age, sex, education, region and the Charlson comorbidity index, whereas our study additionally adjusted for more lifestyle factors. Third, they did not explore specific diseases in each mortality category. Most importantly, their analysis was restricted to MASLD, whereas our study investigated the full spectrum of SLD subtypes, allowing us to directly compare MASLD, MetALD and ALD.

Given the broader range of disease-specific mortality seen in this study, multidisciplinary care is required to address the complex needs of people living with SLD. Since extrahepatic cancer and circulatory disease are the predominating causes of mortality in SLD population, particularly in those with low fibrosis stage, clinical management should prioritise prevention and treatment of these conditions alongside efforts to halt fibrosis progression to reduce overall mortality. Some SLD-related extrahepatic cancers (e.g., colorectal cancer, breast cancer, prostate cancer) could be identified by screening programmes ^25^; however, it remains unclear whether enhanced cancer screening would be cost-effective in population with liver steatosis and how best to implement them. CVD risk assessment is also a critical component of SLD care, as both steatosis and fibrosis are independent risk factors for CVD incidence and mortality ^9,26^.

This study offers several strengths, notably its long follow-up period, large cohort size, and a comprehensive examination of mortality outcomes across ICD10 categories. Nonetheless, there are some limitations. First, liver steatosis and fibrosis were assessed using non-invasive biomarkers of FLI and FIB4, rather than more accurate imaging or biopsy-based methods. While this may have introduced misclassification bias, these scores are widely validated and used in previous research and outperformed liver biopsy in certain populations ^27^. A sensitivity analysis of using HSI replacing FLI generated consistent results, reinforcing the robustness of the main findings. Second, UK Biobank is not fully representative of the wider UK population regarding demography, ethnicity, socioeconomic status and disease prevalence ^28^, which may affect the generalisability of our findings.

To conclude, in this large UK study, SLD was linked to elevated mortality from neoplasm, CVD, digestive disease, chronic kidney disease, diabetes and Covid-19. Among these, CVD and neoplasms were the leading contributors to excess mortality. These findings highlight the wide-ranging systemic impact of SLD beyond liver-related complications, and emphasize the importance of a comprehensive multidisciplinary management strategy that integrates cardiometabolic burden management, cancer prevention, and fibrosis mitigation.

## Data Availability

All data produced in the present study are available upon reasonable request to the authors

## Statements and Declarations

### Author contribution

QF conceived the research idea and conducted data analysis. MW provided data access. QF, MW, PM and CI interpreted results. QF drafted the manuscript. QF, MW, PM and CI critically reviewed and revised the manuscript. QF is the guarantor. The corresponding author attests that all listed authors meet authorship criteria and that no others meeting the criteria have been omitted.

### Ethical approval

UK Biobank has approval from the North West Multi-centre Research Ethics Committee (MREC) as a Research Tissue Bank (RTB) approval. This approval means that researchers do not require separate ethical clearance and can operate under the RTB approval.

### Transparency statement

Q Feng confirms that the manuscript is an honest, accurate, and transparent account of the study being reported; that no important aspects of the study have been omitted; and that any discrepancies from the study as originally planned (and, if relevant, registered) have been explained.

### Data sharing statement

UK Biobank data are available to registered researchers at https://www.ukbiobank.ac.uk/.

## Acknowledgement

We sincerely thank the UK Biobank participants and staff for their contribution to this valuable data resource. This study was conducted under application number 74018.

QF was funded/supported by the NIHR Imperial Biomedical Research Centre (BRC) [NIHR203323]. The views expressed are those of the authors and not necessarily those of the NIHR or the Department of Health and Social Care.

The Section of Investigative Medicine and Endocrinology at Imperial College London is funded by grants from the MRC, NIHR and is supported by the NIHR Biomedical Research Centre Funding Scheme and the NIHR/Imperial Clinical Research Facility. CI is funded by an NIHR Senior Clinical and Practitioner Research Award (NIHR304591) and an NIHR Imperial BRC Pilot Grant (PSR328).

The Division of Digestive Diseases at Imperial College London receives financial support from the National Institute of Health Research (NIHR) Imperial Biomedical Research Centre (BRC) based at Imperial College London and Imperial College Healthcare NHS Trust.

